# Parent-reported assessment scores reflect ASD severity in 2- to 7- year-old children

**DOI:** 10.1101/2022.04.14.22273879

**Authors:** Priyanka Jagadeesan, Adam Kabbani, Andrey Vyshedskiy

**Affiliations:** Boston University, Boston, MA 02215, USA; ImagiRation, Boston, MA 02135, USA

**Keywords:** autism, ASD, language delay, receptive language, language comprehension, combinatorial language, recursive language, prefrontal synthesis, syntactic language

## Abstract

We investigated the relationship between parent-reported assessments and autism spectrum disorder (ASD) severity. Parents evaluated 9573 children with ASD on five subscales: combinatorial receptive language, expressive language, sociability, sensory awareness, and health using Autism Treatment Evaluation Checklist (ATEC) and Mental Synthesis Evaluation Checklist (MSEC). Scores in every subscale improved with age and there were clear differences between the three diagnostic categories. The differences between mild and moderate ASD as well as between moderate and severe ASD reached statistical significance in each subscale and in every age group in children 3 years of age and older. These findings demonstrate a consistent relationship between children’s diagnoses and their assessments.

## Introduction

Clinical trials routinely use parent-reported assessments of children as an outcome measure (Bellini, 2004; Sikich et al., 2021; Tarver et al., 2019; Wood et al., 2009). Parents’ assessments provide additional insights into the course of disease without imposing the extra cost associated with clinicians’ assessments. However, little data exists on the fidelity of parent-report evaluations (Eiser & Morse, 2001). In 2015 we published a language training app for children (Dunn, Elgart, Lokshina, Faisman, Khokhlovich, et al., 2017b, 2017a; Dunn, Elgart, Lokshina, Faisman, Waslick, et al., 2017; Vyshedskiy, Khokhlovich, et al., 2020; Vyshedskiy & Dunn, 2015), which invites parents to complete their child’s evaluation and diagnosis every three months. Parents complete Autism Treatment Evaluation Checklist (ATEC) (Rimland & Edelson, 1999) and Mental Synthesis Evaluation Checklist (MSEC) (Braverman et al., 2018) that assess children along five subscales: combinatorial receptive language, expressive language, sociability, sensory awareness, and health. As a result, we accumulated over a hundred thousand assessments.

Analysis of these parents’ assessments yielded several important insights into the effects of culture and physical conditions on the developmental trajectories of children with ASD. A 3-year study investigating the effect of passive video and television watching in children with ASD (N= 3227) demonstrated that longer video and television watching were associated with a 1.3-fold (*p*=0.0719) faster improvement in development of expressive language, but also resulted in a 1.4-fold (*p*=0.0128) slower development of combinatorial receptive language. The differences in sociability, sensory awareness, and health scores remained insignificant (Fridberg et al., 2021). Similarly, another 3-year study looking at the effect of pretend play (N= 7069) showed that pretend play was associated with a 1.9-fold faster improvement in combinatorial receptive language (*p*<0.0001), a 1.4-fold faster improvement in expressive language (*p*<0.0001), and a 1.3-fold faster improvement in sensory awareness (*p*=0.0009); however, the effect on sociability and health was insignificant. In terms of health studies: seizures and sleep have been analyzed for their impact on developmental trajectories in children with ASD. An analysis of the effect of seizures (N=8461) showed that children with no seizures improved their expressive language 1.3-times faster (*p=*0.0037); sociability 2.3-times faster (*p=*0.0320); sensory awareness 6.2-times faster (*p*=0.0047); and health 20.0-times faster (*p*<0.0001) whereas, the effect on receptive language was insignificant (Forman et al., 2022). Additionally, an investigation of the effect of sleep problems (N=7069) showed that children with no sleep problems improved their sociability 3-times faster (*p*=0.0426) and their health significantly faster (*p*<0.0001; the exact ratio could not be calculated as the health score in children with sleep problems has declined compared to the baseline); the effect on receptive language, expressive language, and sensory awareness was insignificant (Levin et al., 2022). Finally, in a 3-year study of 6454 children, children who engaged with a specialized language therapy displayed a 2.2-fold faster improvement in their combinatorial language scores when compared to children with similar initial evaluations (*p*<0.0001) and 1.4-fold faster improvement in their expressive language score (*p*=0.0144). However, differences in their sociability, sensory awareness, and health scores remained insignificant (Vyshedskiy, DuBois, et al., 2020).

Though these results provide interesting correlations on the impacts of multifactorial cultural and physiological conditions on ASD development, there remains resistance amongst researchers in accepting parent-reported evaluations. There is an understanding within the psychological community that parents can yield to wishful thinking and therefore cannot be trusted with an evaluation of their own children (Scattone et al., 2011). To provide clarity on the fidelity of parent-reports, we have investigated the relationship between children evaluation scores and their ASD severity. The Diagnostic and Statistical Manual of Mental Disorders, 5th Edition (DSM-5) specifies three levels of ASD, depending on severity of disorder and support required in daily life (A. P. Association, 2013). We hypothesized that if parents clearly understood and honestly communicated their child’s diagnosis, the reported ASD severity level would have a consistent relationship with assessment subscales. Wherein, greater ASD severity would correspond to worse assessment scores and vice versa. Conversely, if parents misreported their children’s diagnosis, no difference in the average assessment score would be expected between the groups.

The cross-sectional analysis of 9573 children has demonstrated statistically significant differences between mild and moderate ASD diagnosis as well as between moderate and severe ASD diagnosis in each subscale and in every age group in children 3 years of age and older. These findings are consistent with high fidelity of parent-reported evaluations and their children’s diagnoses.

## Methods

### Participants

Participants were 9573 users of a language therapy app, that was made available gratis at all major app stores in September 2015. Once the app was downloaded, caregivers were asked to register and provide their demographic details, including the child’s diagnosis and age. Participants were asked to select a single diagnosis from the following list: Suspected Autism, Mild Autism, Moderate Autism, Severe Autism, Pervasive Developmental Disorder, Lost Diagnosis of Autism or PDD, Asperger Syndrome, Social Communication Disorder, Specific Language Impairment, Apraxia, Sensory Processing Disorder, Down Syndrome, Other Genetic Disorder, Mild Language Delay, ADHD or ADD, and Normally developed child. This study included participants who selected Mild Autism, Moderate Autism, Severe Autism, or Asperger Syndrome as their child’s diagnosis. The Asperger Syndrome group was combined with the Mild Autism group as per DSM-5 recommendations. Table 1 describes group size and gender information.

**Table 1.**
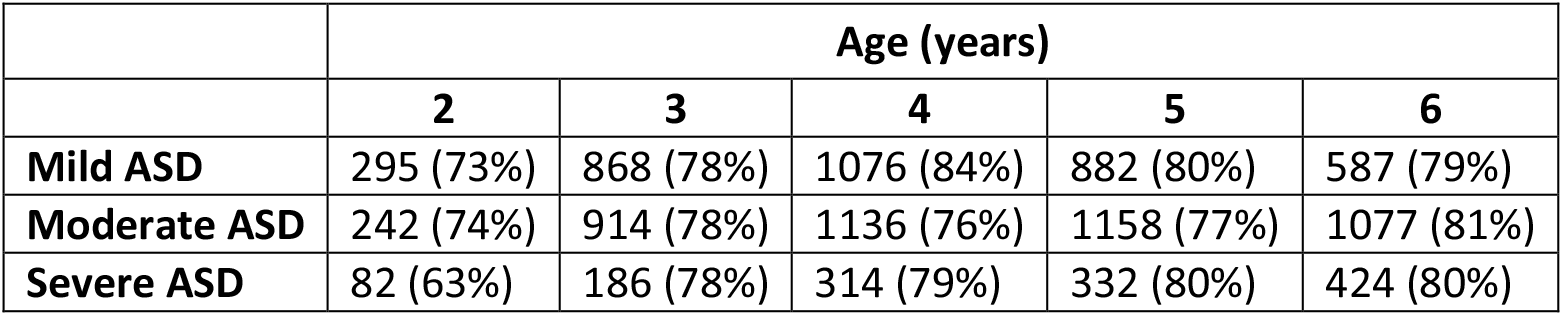
The number of participants in each age group and ASD level. The number in brackets shows percent of males.

|  | Age (years) |  |  |  |  |
| --- | --- | --- | --- | --- | --- |
|  | 2 | 3 | 4 | 5 | 6 |
| **Mild ASD** | 295 (73%) | 868 (78%) | 1076 (84%) | 882 (80%) | 587 (79%) |
| **Moderate ASD** | 242 (74%) | 914 (78%) | 1136 (76%) | 1158 (77%) | 1077 (81%) |
| **Severe ASD** | 82 (63%) | 186 (78%) | 314 (79%) | 332 (80%) | 424 (80%) |

The education level of the participants’ caregivers was as follows: 94% had at least a high school diploma, 73% had at least college education, 35% at least a master’s, and 5% had a doctorate. All data presented in this report is cross-sectional.

### Assessments

Caregivers consented to anonymized data analysis and completed the Mental Synthesis Evaluation Checklist (MSEC) (Braverman et al., 2018) to assess combinatorial receptive language and the Autism Treatment Evaluation Checklist (ATEC) (Rimland & Edelson, 1999) to assess children development. These checklists evaluate development on four domains: expressive language, sociability, sensory awareness, and health. The ATEC questionnaire is comprised of four subscales: 1) Speech/Language/Communication, 2) Sociability, 3) Sensory/Sensory awareness, and 4) Physical/Health/Behavior. The first subscale, Speech/Language/Communication, contains 14 items and its score ranges from 0 to 28 points. The Sociability subscale contains 20 items within a score range of 0 to 40 points. The third subscale, referred here as the Sensory awareness subscale, has 18 items and scores range from 0 to 36 points. The fourth subscale, referred here as the health subscale contains 25 items and scores range from 0 to 75 points. The scores from each subscale are combined to calculate a Total Score, which ranges from 0 to 179 points. A lower score indicates lower severity of ASD symptoms, and a higher score indicates greater severity symptoms of ASD.

### Combinatorial receptive language assessment

The MSEC evaluation was designed complementary to the ATEC in measuring combinatorial receptive language. Out of the 20 MSEC items, those that directly assess receptive language are as following: 1) Understands simple stories that are read aloud; 2) Understands elaborate fairy tales that are read aloud (i.e., stories describing FANTASY creatures); 3) Understands some simple modifiers (i.e., green apple vs. red apple or big apple vs. small apple); 4) Understands several modifiers in a sentence (i.e., small green apple); 5) Understands size (can select the largest/smallest object out of a collection of objects); 6) Understands possessive pronouns (i.e. your apple vs. her apple); 7) Understands spatial prepositions (i.e., put the apple ON TOP of the box vs. INSIDE the box vs. BEHIND the box); 8) Understands verb tenses (i.e., I will eat an apple vs. I ate an apple); 9) Understands the change in meaning when the order of words is changed (i.e., understands the difference between ‘a cat ate a mouse’ vs. ‘a mouse ate a cat’); 10) Understands explanations about people, objects or situations beyond the immediate surroundings (e.g., “Mom is walking the dog,” “The snow has turned to water”). MSEC consists of 20 questions within a score range of 0 to 40 points. Similar to the ATEC, a lower MSEC score indicates a better developed receptive language.

The psychometric quality of MSEC was tested with 3,715 parents of ASD children (Braverman et al., 2018). The Internal reliability of MSEC was good (Cronbach’s alpha > 0.9). Additionally, MSEC also exhibited adequate test–retest reliability, good construct validity, and good known group validity as reflected by the difference in MSEC scores for children of different ASD severity levels. MSEC norms are reported in Ref. (Arnold & Vyshedskiy, 2022).

To simplify interpretation of figure labels, the subscale 1 of the ATEC evaluation is herein referred to as the Expressive Language subscale and the MSEC scale is referred to as the Receptive Language subscale.

## Results

The participants were 9573 parents of children with ASD ranging from 2 to 7 years old. They identified their children’s diagnosis as mild, moderate, or severe ASD and assessed their children on five subscales: combinatorial receptive language, expressive language, sociability, sensory awareness, and health. The mild ASD group showed superior scores than the moderate ASD group, and the moderate ASD group had better scores than the severe ASD group in all subscales across all ages as (Ref. to Figure 1).

**Figure 1.**
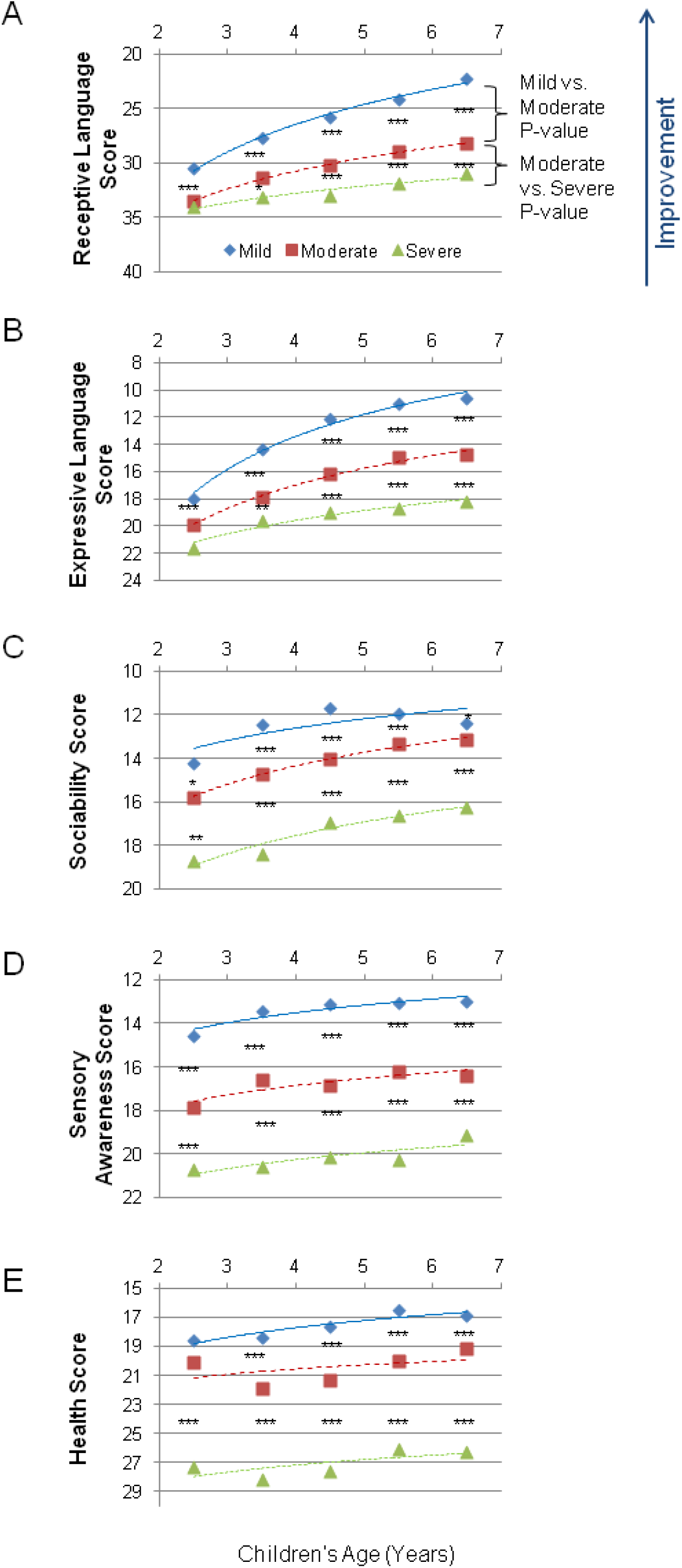
Averaged scores in five subscales for the mild, moderate, and severe ASD. A lower score indicates symptom improvement. P-value is marked: ***<0.0001; **<0.001; *<0.05. (A) Receptive Language score. (B) Expressive Language score. (C) Sociability score. (D) Cognitive awareness score (E) Health score.

On the Receptive Language subscale (Table 2), the difference between the mild ASD group and the moderate ASD group was statistically significant in children 3 years of age (0.008) as well as in age groups 4, 5, and 6 years of age (*p*<0.0001). However, the difference had not reached statistical significance in 2 years of age group (*p*=0.53). In contrast, the difference between the moderate ASD group and the severe ASD group was statistically significant (*p*<0.0001) in every age group studied.

**Table 2.** Averaged Combinatorial Receptive Language score (assessed by MSEC) for the mild, moderate, and severe ASD. A lower score indicates symptom improvement. Data are presented as Mean(SD).

|  | Age (years) |  |  |  |  |
| --- | --- | --- | --- | --- | --- |
|  | 2 | 3 | 4 | 5 | 6 |
| **Mild ASD** | 30.4 (7.5) | 27.7 (7.9) | 25.8 (7.7) | 24.1 (8.4) | 22.3 (9.2) |
| **Moderate ASD** | 33.5 (5.9) | 31.4 (6.8) | 30.1 (7.3) | 28.9 (7.5) | 28.2 (7.8) |
| **Severe ASD** | 34 (8) | 33.1 (8) | 33 (6.8) | 31.9 (7.5) | 31 (7.9) |

The Expressive Language subscale (Table 3) showed a similar trend to scores for receptive language, with the difference between the mild ASD group and the moderate ASD group being statistically significant in children 3 years of age (0.008) as well as in age groups 4, 5, and 6 years of age (*p*<0.0001) but not in 2 years of age group (*p*=0.46). Similarly, the difference between the moderate ASD group and the severe ASD group was statistically significant (*p*<0.0001) in every age group.

**Table 3.** Averaged Expressive Language (assessed by ATEC subscale 1) score for the mild, moderate, and severe ASD. A lower score indicates symptom improvement. Data are presented as Mean(SD).

|  | Age (years) |  |  |  |  |
| --- | --- | --- | --- | --- | --- |
|  | 2 | 3 | 4 | 5 | 6 |
| **Mild ASD** | 18 (5.9) | 14.4 (6.7) | 12.1 (6.5) | 11 (6.2) | 10.6 (6.2) |
| **Moderate ASD** | 20 (5.2) | 17.8 (5.8) | 16.2 (6.2) | 14.9 (6.1) | 14.7 (5.7) |
| **Severe ASD** | 21.6 (5.3) | 19.6 (5.6) | 19 (5.6) | 18.7 (5.6) | 18.1 (5.7) |

On the Sociability subscale (Table 4), the difference between the mild ASD group and the moderate ASD group was statistically significant in children 2 years of age (0.012) and in age groups 3, 4, 5, and 6 years of age (*p*<0.0001). The difference between the moderate ASD group and the severe ASD group was statistically significant in children 2 years of age (0.0004), as well as in groups 3, 4, 5, and 6 years of age (*p*<0.0001).

**Table 4.** Averaged Sociability (assessed by ATEC subscale 2) score for the mild, moderate, and severe ASD. A lower score indicates symptom improvement. Data are presented as Mean(SD).

|  | Age (years) |  |  |  |  |
| --- | --- | --- | --- | --- | --- |
|  | 2 | 3 | 4 | 5 | 6 |
| **Mild ASD** | 14.2 (7.3) | 12.4 (7.5) | 11.7 (7.3) | 11.9 (7.5) | 12.3 (7.3) |
| **Moderate ASD** | 15.8 (7.3) | 14.7 (7.5) | 14 (7.2) | 13.3 (6.9) | 13.1 (6.9) |
| **Severe ASD** | 18.7 (7.9) | 18.4 (7.7) | 16.9 (7.8) | 16.6 (8.1) | 16.3 (7.7) |

On the Sensory awareness subscale (Table 5), the differences between the mild ASD group and the moderate ASD group was statistically significant in every age group (*p*<0.0001). The difference between the moderate ASD group and the severe ASD group was also statistically significant in every age group (*p*<0.0001).

**Table 5.** Averaged Sensory Awareness (assessed by ATEC subscale 3) score for the mild, moderate, and severe ASD. A lower score indicates symptom improvement. Data are presented as Mean(SD).

|  | Age (years) |  |  |  |  |
| --- | --- | --- | --- | --- | --- |
|  | 2 | 3 | 4 | 5 | 6 |
| **Mild ASD** | 14.6 (6.1) | 13.4 (6.5) | 13.1 (6.4) | 13.1 (6.8) | 13 (6.5) |
| **Moderate ASD** | 17.9 (5.8) | 16.6 (6.4) | 16.8 (6.6) | 16.2 (6.1) | 16.4 (6.3) |
| **Severe ASD** | 20.7 (6.4) | 20.6 (6.7) | 20.2 (6.2) | 20.3 (6.2) | 19.1 (6.5) |

On the Health subscale (Table 6), the difference between the mild ASD group and the moderate ASD group was statistically significant in children in every age group (*p*<0.0001). The difference between the moderate ASD group and the severe ASD group had not reached statistical significance in 2 years of age group (*p*=0.526) but was statistically significant (*p*<0.0001) in older children.

**Table 6.** Averaged Health (assessed by ATEC subscale 4) score for the mild, moderate, and severe ASD. A lower score indicates symptom improvement. Data are presented as Mean(SD).

|  | Age (years) |  |  |  |  |
| --- | --- | --- | --- | --- | --- |
|  | 2 | 3 | 4 | 5 | 6 |
| **Mild ASD** | 18.6 (11.8) | 18.4 (7.9) | 17.6 (11.3) | 16.5 (10.9) | 16.9 (10.5) |
| **Moderate ASD** | 20.1 (11.8) | 21.9 (12.4) | 21.4 (12.2) | 19.9 (11.2) | 19.2 (10.5) |
| **Severe ASD** | 27.3 (15.1) | 28.2 (13.9) | 27.6 (12.7) | 26 (12.8) | 26.3 (13.5) |

Figure 2 shows the combined measures: the Total ATEC score (Table 7) and the MSEC score + the Total ATEC score (Table 8). In both combined measures, the difference between the mild ASD group and the moderate ASD group as well as between the moderate ASD group and the severe ASD group was statistically significant in children in every age group (*p*<0.0001).

**Table 7.** Averaged Total ATEC score for the mild, moderate, and severe ASD. A lower score indicates symptom improvement. Data are presented as Mean(SD).

|  | Age (years) |  |  |  |  |
| --- | --- | --- | --- | --- | --- |
|  | 2 | 3 | 4 | 5 | 6 |
| **Mild ASD** | 65.6 (21.3) | 58.7 (23.3) | 54.5 (23.1) | 52.5 (23) | 52.8 (22.4) |
| **Moderate ASD** | 73.9 (20.6) | 71.1 (23.5) | 68.4 (24.3) | 64.5 (21.9) | 63.4 (21.2) |
| **Severe ASD** | 88.4 (23.9) | 86.9 (25.6) | 83.9 (23.4) | 81.8 (24.1) | 79.8 (23.7) |

**Table 8.** Averaged Total ATEC + MSEC score for the mild, moderate, and severe ASD. A lower score indicates symptom improvement. Data are presented as Mean(SD).

|  | Age (years) |  |  |  |  |
| --- | --- | --- | --- | --- | --- |
|  | 2 | 3 | 4 | 5 | 6 |
| **Mild ASD** | 96 (23.6) | 86.4 (27.9) | 80.3 (27.2) | 76.6 (27.7) | 75.1 (27.7) |
| **Moderate ASD** | 107.4 (23) | 102.5 (26.6) | 98.6 (28.2) | 93.4 (26.1) | 91.7 (25.6) |
| **Severe ASD** | 122.4 (25.2) | 120 (28.8) | 116.9 (25.8) | 113.6 (26.6) | 110.8 (27.5) |

**Figure 2.**
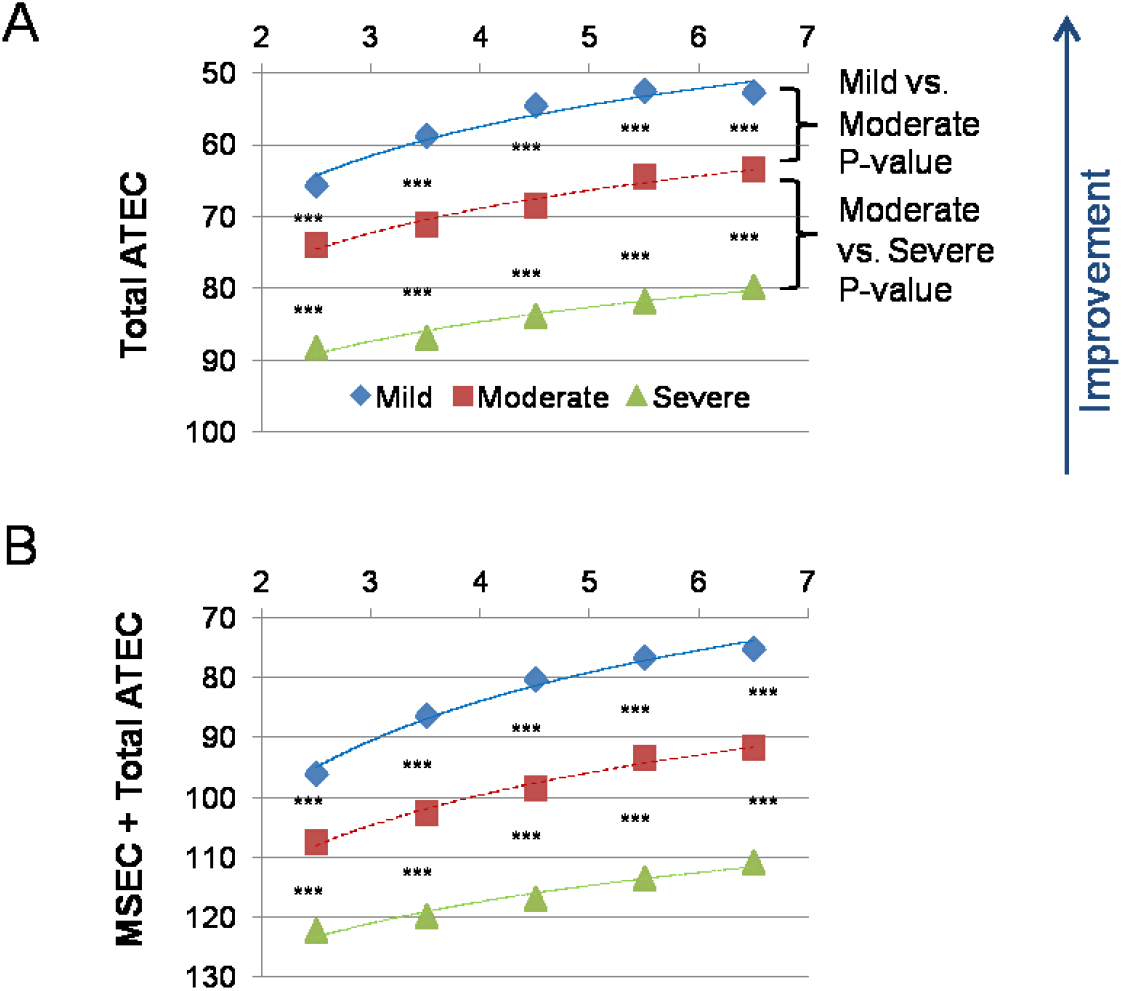
Averaged scores in five subscales for the mild, moderate, and severe ASD. A lower score indicates symptom improvement. (A) The total ATEC score, which is the sum of Expressive Language score, Sociability score, Cognitive awareness score, and Health score. (B) The sum of the total ATEC and the MSEC score (i.e. the sum of all five orthogonal subscales). A lower score indicates symptom improvement. P-value is marked: ***<0.0001.

## Discussion

This study evaluates the fidelity of parents’ assessments by comparing parent-reported evaluation scores with children’s ASD severity. Despite the common use of parent-report assessments in clinical trials (Bellini, 2004; Sikich et al., 2021; Tarver et al., 2019; Wood et al., 2009), there is little research evaluating their reliability (Scattone et al., 2011). Since we had access to both parent-reported assessments and children’s diagnosis, we had the opportunity to compare these scores between diagnoses. To our satisfaction, we have found significant differences between diagnostic categories. As expected, children diagnosed with mild ASD had better scores than children with moderate ASD, and children diagnosed with moderate ASD had better scores than those with severe ASD. The differences reached statistical significance in each subscale and in every age group for children 3 years and older as well as, in the cumulative Total ATEC and Total ATEC+MSEC scales for children 2 years of age and older. These findings suggest high fidelity of parents’ assessments.

It’s crucial to note that the ATEC is not a diagnostic checklist and was initially designed to evaluate the effectiveness of treatment (Rimland & Edelson, 1999). Therefore, ASD severity can only have an approximate relationship with the total ATEC score and age. Table 9 and 10 list approximated ATEC total scores and approximated MSEC+ATEC total scores as they relate to ASD severity and age, respectively.

**Table 9.** Approximate relationship between ATEC total score, age, and ASD severity. At any age, a greater ATEC score indicates greater ASD severity.

|  | Age (years) |  |  |  |  |
| --- | --- | --- | --- | --- | --- |
|  | 2 | 3 | 4 | 5 | 6 |
| **Mild ASD** | <70 | <65 | <58 | <58 | <58 |
| **Moderate ASD** | 70-81 | 65-79 | 58-76 | 58-73 | 58-72 |
| **Severe ASD** | 81-179 | 79-179 | 76-179 | 73-179 | 72-179 |

**Table 10.** Approximate relationship between MSEC+ATEC total score, age, and ASD severity. At any age, a greater score indicates greater ASD severity.

|  | Age (years) |  |  |  |  |
| --- | --- | --- | --- | --- | --- |
|  | 2 | 3 | 4 | 5 | 6 |
| **Mild ASD** | <102 | <94 | <89 | <85 | <83 |
| **Moderate ASD** | 102-115 | 94-111 | 89-108 | 85-104 | 83-101 |
| **Severe ASD** | 115-209 | 111-209 | 108-209 | 104-209 | 101-209 |

On the Receptive Language subscale, significant differences were detected between mild and moderate ASD at 2 years of age (*p*<0.0001, Figure 1A). This observation combined with the observation of a significant difference between MSEC scores comparing neurotypical and ASD children by Arnold et al. (Arnold & Vyshedskiy, 2022) suggests that the assessment of combinatorial receptive language holds merit for diagnosing and monitoring of language deficits.

Consistent with previous reports (Brignell et al., 2018), the difference in Receptive and Expressive language scores between ASD levels increased with age with mild ASD showing greater gains compared to moderate and severe ASD (Figure 1. A and B). It’s crucial to note that even mild ASD scores remain significantly behind the normal range (compare mild ASD receptive language score of 30.4±7.5 at 2 years of age to the score of 22.8±4.2 observed in neurotypical children at the same age; and at 6 years of age, the score of 22.3±9.2 versus 4.5±2.4) (Arnold & Vyshedskiy, 2022). Conversely, the difference in Sociability, Sensory Awareness, and Health score between ASD levels decreased or remained unchanged with age (Figure 1. C to E).

While limited in its range of application across disorders, this study contributes to affirming the reliability of parents’ assessments in evaluating their children with ASD and provides additional evidence in support of fidelity of such evaluations for autism spectrum disorder (Ebert, 2017). To our knowledge this is the first investigation of relationship between parent-report ATEC/MSEC score and ASD severity for the range of 2 to 7 year old children. Future studies may compare parent-report instruments such as ATEC and MSEC to clinicians’ assessments, such as Childhood Autism Rating Scale (CARS) (Schopler et al., 2002) and Autism Diagnostic Observation Schedule (ADOS) (Gotham et al., 2009), in order to further explore the reliability of parents’ evaluations.

## Data Availability

All data produced in the present study are available upon reasonable request to the authors

## Acknowledgements

We wish to thank Dr. Petr Ilyinskii for his scrupulous editing of this manuscript.

## Author contributions

AV designed the study. AV and AK analyzed the data. AV and PJ wrote the paper.

## Informed Consent

Caregivers have consented to anonymized data analysis and publication of the results. The study was conducted in compliance with the Declaration of Helsinki (W. M. Association, 2013).

## Compliance with Ethical Standards

Using the Department of Health and Human Services regulations found at 45 CFR 46.101(b)(4), the Biomedical Research Alliance of New York LLC Institutional Review Board (IRB) determined that this research project is exempt from IRB oversight.

## Data Availability

De-identified raw data from this manuscript are available from the corresponding author upon reasonable request.

## Code availability statement

Code is available from the corresponding author upon reasonable request.

## References

Arnold, M., & Vyshedskiy, A. (2022). Combinatorial language parent-report score differs significantly between typically developing children and those with Autism Spectrum Disorders. MedRxiv, 2022.03.30.22271503. https://doi.org/10.1101/2022.03.30.22271503

Association, A. P. (2013). Diagnostic and statistical manual of mental disorders (DSM-5®). American Psychiatric Pub.

Association, W. M. (2013). World Medical Association Declaration of Helsinki: Ethical principles for medical research involving human subjects. Jama, 310(20), 2191–2194.

Bellini, S. (2004). Social skill deficits and anxiety in high-functioning adolescents with autism spectrum disorders. Focus on Autism and Other Developmental Disabilities, 19(2), 78–86.

Braverman, J., Dunn, R., & Vyshedskiy, A. (2018). Development of the Mental Synthesis Evaluation Checklist (MSEC): A Parent-Report Tool for Mental Synthesis Ability Assessment in Children with Language Delay. Children, 5(5), 62. https://doi.org/10.3390/children5050062

Brignell, A., Morgan, A. T., Woolfenden, S., Klopper, F., May, T., Sarkozy, V., & Williams, K. (2018). A systematic review and meta-analysis of the prognosis of language outcomes for individuals with autism spectrum disorder. Autism & Developmental Language Impairments, 3, 2396941518767610.

Dunn, R., Elgart, J., Lokshina, L., Faisman, A., Khokhlovich, E., Gankin, Y., & Vyshedskiy, A. (2017a). Children With Autism Appear To Benefit From Parent-Administered Computerized Cognitive And Language Exercises Independent Of the Child’s Age Or Autism Severity. Autism Open Access, 7(217). https://doi.org/10.4172/2165-7890.1000217

Dunn, R., Elgart, J., Lokshina, L., Faisman, A., Khokhlovich, E., Gankin, Y., & Vyshedskiy, A. (2017b). Comparison of performance on verbal and nonverbal multiple-cue responding tasks in children with ASD. Autism Open Access, 7, 218. https://doi.org/10.4172/2165-7890.1000218

Dunn, R., Elgart, J., Lokshina, L., Faisman, A., Waslick, M., Gankin, Y., & Vyshedskiy, A. (2017). Tablet-Based Cognitive Exercises as an Early Parent-Administered Intervention Tool for Toddlers with Autism—Evidence from a Field Study. Clinical Psychiatry, 3(1). https://doi.org/10.21767/2471-9854.100037

Ebert, K. D. (2017). Convergence between parent report and direct assessment of language and attention in culturally and linguistically diverse children. Plos One, 12(7), e0180598.

Eiser, C., & Morse, R. (2001). Quality-of-life measures in chronic diseases of childhood. Health Technology Assessment (Winchester, England), 5(4), 1–157.

Forman, P., Edward, E., & Vyshedskiy, A. (2022). Effect of seizures on developmental trajectories in children with autism. MedRxiv, 2022.03.30.22273179. https://doi.org/10.1101/2022.03.30.22273179

Fridberg, E., Khokhlovich, E., & Vyshedskiy, A. (2021). Watching Videos and Television Is Related to a Lower Development of Complex Language Comprehension in Young Children with Autism. Healthcare, 9(4), 423.

Gotham, K., Pickles, A., & Lord, C. (2009). Standardizing ADOS scores for a measure of severity in autism spectrum disorders. Journal of Autism and Developmental Disorders, 39(5), 693–705.

Levin, J., Khokhlovich, E., & Vyshedskiy, A. (2022). Sleep problems effect on developmental trajectories in children with autism. MedRxiv, 2022.03.30.22273178. https://doi.org/10.1101/2022.03.30.22273178

Rimland, B., & Edelson, S. (1999). Autism Research Institute. Autism Treatment Evaluation Checklist (ATEC).

Scattone, D., Raggio, D. J., & May, W. (2011). Comparison of the vineland adaptive behavior scales, and the bayley scales of infant and toddler development. Psychological Reports, 109(2), 626–634.

Schopler, E., Reichler, R. J., & Renner, B. R. (2002). The childhood autism rating scale (CARS). Western Psychological Services Los Angeles, CA.

Sikich, L., Kolevzon, A., King, B. H., McDougle, C. J., Sanders, K. B., Kim, S.-J., Spanos, M., Chandrasekhar, T., Trelles, M. P., & Rockhill, C. M. (2021). Intranasal Oxytocin in Children and Adolescents with Autism Spectrum Disorder. New England Journal of Medicine, 385(16), 1462–1473.

Tarver, J., Palmer, M., Webb, S., Scott, S., Slonims, V., Simonoff, E., & Charman, T. (2019). Child and parent outcomes following parent interventions for child emotional and behavioral problems in autism spectrum disorders: A systematic review and meta-analysis. Autism, 23(7), 1630–1644.

Vyshedskiy, A., DuBois, M., Mugford, E., Piryatinsky, I., Radi, K., Braverman, J., & Maslova, V. (2020). Novel Linguistic Evaluation of Prefrontal Synthesis (LEPS) test measures prefrontal synthesis acquisition in neurotypical children and predicts high-functioning versus low-functioning class assignment in individuals with autism. Applied Neuropsychology: Child. https://doi.org/10.1080/21622965.2020.1758700

Vyshedskiy, A., & Dunn, R. (2015). Mental Imagery Therapy for Autism (MITA)-An Early Intervention Computerized Brain Training Program for Children with ASD. Autism Open Access, 5(1000153), 2.

Vyshedskiy, A., Khokhlovich, E., Dunn, R., Faisman, A., Elgart, J., Lokshina, L., Gankin, Y., Ostrovsky, S., deTorres, L., & Edelson, S. M. (2020). Novel prefrontal synthesis intervention improves language in children with autism. Healthcare, 8(4), 566. https://doi.org/doi.org/10.3390/healthcare8040566

Wood, J. J., Drahota, A., Sze, K., Van Dyke, M., Decker, K., Fujii, C., Bahng, C., Renno, P., Hwang, W.-C., & Spiker, M. (2009). Brief report: Effects of cognitive behavioral therapy on parent-reported autism symptoms in school-age children with high-functioning autism. Journal of Autism and Developmental Disorders, 39(11), 1608–1612.

